# Rare Genetic Variants Associated with Sudden Cardiac Arrest in the Young: A Prospective, Population-Based Study

**DOI:** 10.1101/2022.10.27.22281332

**Authors:** Lauri Holmstrom, Ninad S Chaudhary, Kotoka Nakamura, Harpriya Chugh, Audrey Uy-Evanado, F Faye Norby, Ginger A Metcalf, Vipin K Menon, Bing Yu, Eric Boerwinkle, Sumeet S Chugh, Zeynep Akdemir, Evan P Kransdorf

## Abstract

**Background:** Sudden cardiac arrest (SCA) is a rare and tragic event among the young and often caused by inherited cardiac disease. Previous studies have investigated referral cohorts, but the prevalence of disease-associated variants is unclear at the community level. We investigated the prevalence of genetic variants among community-based cases of SCA aged <21 years.

**Methods:** The study sample is obtained from two prospective, community-based studies of out-of-hospital SCA ongoing in the Portland, OR metro area (population ∼1 million) and Ventura County CA (population ∼850,000). We performed next-generation whole genome sequencing and then rare variant analysis of candidate genes associated with arrhythmic syndromes and cardiomyopathy in ClinGen.

**Results:** The mean age of the study subjects was 11.3±8.0 (30% non-white, 45% female). We found that 36 of 52 young SCA victims (69%) harbored uncertain, likely pathogenic (LP), or pathogenic (P) variants. Eight subjects (15%) carried 9 LP/P variants. Patients with clinical histories suggesting primary arrhythmic syndromes or hypertrophic cardiomyopathy were more likely to harbor clinically actionable variants or variants of unknown significance (VUS), than subjects with myocarditis, sudden infant death syndrome, or sudden arrhythmic death. Variants were more likely to be classified as LP/P among Whites (8/9, 88.9%) as compared to non-Whites (1/9, 11.1%, p = 0.036).

**Conclusions:** A notable proportion of young SCA victims in the community harbor rare, potentially disease-associated gene variants, and further studies are needed to understand variants of unknown significance. We identified differences by phenotype groups and race that have potential implications for genetic testing.

## Introduction

The societal burden of sudden cardiac arrest (SCA) is high ^1^. An autopsy will reveal a cardiac cause of SCA such as critical CAD in some cases, but many cases remain unexplained ^2^. In unexplained cases, genetic testing may reveal the presence of rare genetic variants in genes previously associated with structural heart disease ^3^.

Children and young adults are less likely to suffer SCA compared to middle-aged and older persons^4, 5^. Autopsy results fail to reveal an explanation for SCA in approximately 40% of cases^6^ and the attributed cause of death may be inaccurate ^7^. Studies using genetic testing have revealed a significant portion (∼12-30%) of children/young adults with sudden cardiac death (SCD) that have pathogenic rare variants in genes associated with arrhythmic syndromes and cardiomyopathy ^6, 8^. In addition, rare variants in genes associated with epilepsy have also been associated with SCA in the young^9^.

Studies on the prevalence of rare gene variants in children and young adults with SCA have largely been performed in referral cohorts with mostly European ancestry and thus the prevalence and impact of rare variants among unselected young SCA victims in a diverse community setting are not known. In the present study, we used data from two community-based studies to perform whole genome sequencing and then analysis of candidate genes for rare variants in 52 young SCA victims.

## Methods

### Study Cohort

Our study subjects are drawn from 2 prospective, community-based studies of SCA in the United States: the Oregon Sudden Unexpected Death Study (Oregon SUDS, since 2002) and the Ventura Prediction of Sudden Death in Multi-Ethnic Communities study (Ventura PRESTO, since 2015). The study protocols have been described earlier in more detail^10, 11^ and only a brief report follows. Both Oregon SUDS and Ventura PRESTO use an identical design, ascertaining all out-of-hospital SCAs from the community: Oregon SUDS from the Portland Oregon metro area (population ∼1 million), and Ventura PRESTO from Ventura County (population ∼850,000). Potential SCD cases are identified via multilevel data collection which includes data from the region’s 2-tiered Emergency Medical Services (EMS) system, regional hospitals, and the state Medical Examiner’s office. Each potential SCD case underwent comprehensive in-house adjudication that was performed by trained physician-researchers. All available data from medical records, medical examiner’s records, the region’s hospital systems, and EMS reports were used to adjudicate each potential SCD case. We defined SCA as a witnessed sudden loss of pulse of likely cardiac etiology, or if the case was unwitnessed, the subject should have been seen alive within 24 h in good health. Survivors of sudden cardiac arrest were included. Cases with terminal illness and cases that were adjudicated to have likely non-cardiac SCA were excluded. Between March 2002 and January 2021, 8,210 SCA cases were ascertained in the two communities. Of these 209 were <21 years of which 52 underwent next-generation whole genome sequencing and were included in the present study. Genomic DNA was extracted from blood samples that were collected by EMS at the time of SCA, during the survivor’s research visit, or obtained at autopsy. The primary cause of death was determined based on the combination of EMS reports, clinical records, and the state medical examiner’s autopsy reports, using contemporary guidelines and definitions for each phenotype. Based on the medical records review, patients were assigned to one of 5 groups: probable primary arrhythmia syndrome, hypertrophic cardiomyopathy (HCM), myocarditis, sudden infant death syndrome (SIDS), and sudden arrhythmic death syndrome (SADS)^12-15^. The study protocol was approved by the institutional review boards of Ventura County Medical Center, Oregon Health and Science University, and the Cedars-Sinai Health System.

### Sequencing

The DNA was sequenced at Baylor Human Genomic Science Center and processed using a validated next-generation sequence in the Mercury analytical pipeline. The read sequences were first mapped to the GRCh38 reference genome via the Burrows-Wheeler Alignment-MEM algorithm to generate a binary alignment/map file (BAM). Using SAMtools, Picard, and GATK in combination, the BAM files were recalibrated for variant calling into compressed columnar file (CRAM) format. Additional details about the Mercury pipeline have been described elsewhere^16^.

### Variant Calling

CRAMs were reprocessed further in the Mercury to call variants and produce a project-level variant file (gVCFs or VCFs) using the Atlas2-SNP and Atlas-indel callers. An individual genotype file for each sample was obtained in this project.

### Candidate Gene Selection

We utilized the Clinical Genome Resource (ClinGen) Gene-Disease Clinical Validity curations for arrhythmic and cardiomyopathic diseases. For arrhythmia, these included 1-Brugada syndrome (BrS) ^17^, 2-catecholaminergic polymorphic ventricular tachycardia (CPVT) ^18^, 3-short QT syndrome (SQTS) ^18^, and 4-long QT syndrome (LQTS) ^19^. For cardiomyopathy, these included 1-arrhythmogenic right ventricular cardiomyopathy (ARVC) ^20^, dilated cardiomyopathy (DCM) ^21^, and hypertrophic cardiomyopathy (HCM) ^22^. We included genes with “definitive”, “moderate”, “limited”, and “disputed” association with these disorders. These genes are their classifications are detailed in Table 1. We added 4 additional genes associated with sudden death in epilepsy (SUDEP): DEPDC5, SCN1A, SCN4A, and SCN8A ^23, 24^.

**Table 1.** Gene-disease associations based on ClinGen. Genes with definitive, moderate, limited, and disputed association were analyzed in the present study.

| Gene | ARVC | BRS | CPVT | DCM | HCM | LQTS | SQTS |
| --- | --- | --- | --- | --- | --- | --- | --- |
| ABCC9 | 0 | 0 | 0 | L | 0 | 0 | 0 |
| ACTC1 | NKDR | 0 | 0 | M | D | 0 | 0 |
| ACTN2 | 0 | 0 | 0 | 0 | M | 0 | 0 |
| AKAP9 | 0 | 0 | 0 | 0 | 0 | Dis | 0 |
| ANK2 | 0 | Dis | Dis | 0 | 0 | 0 | 0 |
| ANKRD1 | 0 | 0 | 0 | L | L | 0 | 0 |
| BAG3 | 0 | 0 | 0 | D | 0 | 0 | 0 |
| CACNA1C | 0 | Dis | 0 | 0 | 0 | M | Dis |
| CACNA2D1 | 0 | Dis | 0 | 0 | 0 | 0 | Dis |
| CACNB2 | 0 | Dis | 0 | 0 | 0 | 0 | Dis |
| CALM1 | 0 | 0 | M | 0 | 0 | D | 0 |
| CALM2 | 0 | 0 | M | 0 | 0 | D | 0 |
| CALM3 | 0 | 0 | M | 0 | 0 | D | 0 |
| CALR3 | 0 | 0 | 0 | 0 | L | 0 | 0 |
| CASQ2 | 0 | 0 | M | 0 | 0 | 0 | 0 |
| CDH2 | L | 0 | 0 | 0 | 0 | 0 | 0 |
| CSRP3 | 0 | 0 | 0 | L | M | 0 | 0 |
| CTF1 | 0 | 0 | 0 | L | 0 | 0 | 0 |
| CTNNA3 | L | 0 | 0 | 0 | 0 | 0 | 0 |
| DES | M | 0 | 0 | D | 0 | 0 | 0 |
| DSC2 | D | 0 | 0 | 0 | 0 | 0 | 0 |
| DSG2 | D | 0 | 0 | L | 0 | 0 | 0 |
| DSP | D | 0 | 0 | 0 | 0 | 0 | 0 |
| DTNA | 0 | 0 | 0 | L | 0 | 0 | 0 |
| EYA4 | 0 | 0 | 0 | L | 0 | 0 | 0 |
| FLNC | 0 | 0 | 0 | D | D | 0 | 0 |
| FXN | 0 | 0 | 0 | 0 | D | 0 | 0 |
| GATAD1 | 0 | 0 | 0 | L | 0 | 0 | 0 |
| GPD1L | 0 | Dis | 0 | 0 | 0 | 0 | 0 |
| HCN4 | 0 | Dis | 0 | 0 | 0 | 0 | 0 |
| ILK | 0 | 0 | 0 | L | 0 | 0 | 0 |
| JPH2 | 0 | 0 | 0 | M | M | 0 | 0 |
| KCND3 | 0 | Dis | 0 | 0 | 0 | 0 | 0 |
| KCNE1 | 0 | 0 | 0 | 0 | 0 | L | 0 |
| KCNE2 | 0 | 0 | 0 | 0 | 0 | Dis | 0 |
| KCNE3 | 0 | Dis | 0 | 0 | 0 | 0 | 0 |
| KCNE5 | 0 | Dis | 0 | 0 | 0 | 0 | 0 |
| KCNH2 | 0 | Dis | 0 | 0 | 0 | D | D |
| KCNJ2 | 0 | 0 | Dis | 0 | 0 | L | M |
| KCNJ5 | 0 | 0 | 0 | 0 | 0 | Dis | 0 |
| KCNJ8 | 0 | Dis | 0 | 0 | 0 | 0 | 0 |
| KCNQ1 | 0 | 0 | 0 | 0 | 0 | D | S |
| KLF10 | 0 | 0 | 0 | 0 | L | 0 | 0 |
| LAMA4 | 0 | 0 | 0 | L | 0 | 0 | 0 |
| LAMP2 | 0 | 0 | 0 | 0 | D | 0 | 0 |
| LDB3 | Dis | 0 | 0 | L | 0 | 0 | 0 |
| LMNA | L | 0 | 0 | D | 0 | 0 | 0 |
| MYBPC3 | L | 0 | 0 | L | D | 0 | 0 |
| MYH6 | 0 | 0 | 0 | L | L | 0 | 0 |
| MYH7 | L | 0 | 0 | D | D | 0 | 0 |
| MYL2 | NKDR | 0 | 0 | L | D | 0 | 0 |
| MYL3 | L | 0 | 0 | Dis | D | 0 | 0 |
| MYLK2 | 0 | 0 | 0 | 0 | L | 0 | 0 |
| MYOM1 | 0 | 0 | 0 | 0 | L | 0 | 0 |
| MYOZ2 | 0 | 0 | 0 | 0 | L | 0 | 0 |
| MYPN | 0 | 0 | 0 | L | L | 0 | 0 |
| NEBL | 0 | 0 | 0 | L | 0 | 0 | 0 |
| NEXN | 0 | 0 | 0 | M | L | 0 | 0 |
| NKX2-5 | 0 | 0 | 0 | L | 0 | 0 | 0 |
| OBSCN | 0 | 0 | 0 | L | 0 | 0 | 0 |
| PDLIM3 | 0 | 0 | 0 | Dis | L | 0 | 0 |
| PKP2 | D | Dis | Dis | Dis | 0 | 0 | 0 |
| PLEKHM2 | 0 | 0 | 0 | L | 0 | 0 | 0 |
| PLN | M | 0 | 0 | 0 | D | 0 | 0 |
| PRDM16 | 0 | 0 | 0 | L | 0 | 0 | 0 |
| PRKAG2 | 0 | 0 | 0 | 0 | D | 0 | 0 |
| PSEN1 | 0 | 0 | 0 | Dis | 0 | 0 | 0 |
| PSEN2 | 0 | 0 | 0 | L | 0 | 0 | 0 |
| RANGRF | 0 | Dis | 0 | 0 | 0 | 0 | 0 |
| RBM20 | 0 | 0 | 0 | D | 0 | 0 | 0 |
| RYR2 | R | 0 | D | 0 | L | 0 | 0 |
| SCN10A | 0 | Dis | 0 | 0 | 0 | 0 | 0 |
| SCN1B | 0 | Dis | 0 | 0 | 0 | 0 | 0 |
| SCN2B | 0 | Dis | 0 | 0 | 0 | 0 | 0 |
| SCN3B | 0 | Dis | 0 | 0 | 0 | 0 | 0 |
| SCN4B | 0 | 0 | 0 | 0 | 0 | Dis | 0 |
| SCN5A | L | D | Dis | D | 0 | D | Dis |
| SGCD | 0 | 0 | 0 | L | 0 | 0 | 0 |
| SLC22A5 | 0 | 0 | 0 | 0 | 0 | 0 | Dis |
| SLC25A4 | 0 | 0 | 0 | 0 | D | 0 | 0 |
| SLC4A3 | 0 | 0 | 0 | 0 | 0 | 0 | M |
| SLMAP | 0 | Dis | 0 | 0 | 0 | 0 | 0 |
| SNTA1 | 0 | 0 | 0 | 0 | 0 | Dis | 0 |
| TBX20 | 0 | 0 | 0 | L | 0 | 0 | 0 |
| TCAP | 0 | 0 | 0 | L | L | 0 | 0 |
| TECRL | 0 | 0 | D | 0 | 0 | 0 | 0 |
| TGFB3 | L | 0 | 0 | 0 | 0 | 0 | 0 |
| TJP1 | L | 0 | 0 | 0 | 0 | 0 | 0 |
| TMEM43 | D | 0 | 0 | 0 | 0 | 0 | 0 |
| TNNC1 | NKDR | 0 | 0 | D | M | 0 | 0 |
| TNNI3 | NKDR | 0 | 0 | M | D | 0 | 0 |
| TNNI3K | 0 | 0 | 0 | L | 0 | 0 | 0 |
| TNNT2 | NKDR | 0 | 0 | D | D | 0 | 0 |
| TPM1 | NKDR | 0 | 0 | M | D | 0 | 0 |
| TRDN | 0 | 0 | D | 0 | 0 | S | 0 |
| TRIM63 | 0 | 0 | 0 | 0 | L | 0 | 0 |
| TRPM4 | 0 | Dis | 0 | 0 | 0 | 0 | 0 |
| TTN | L | 0 | 0 | D | L | 0 | 0 |
| TTR | 0 | 0 | 0 | 0 | D | 0 | 0 |
| VCL | 0 | 0 | 0 | M | L | 0 | 0 |

Abbreviations: ARVC: arrhythmogenic right ventricular cardiomyopathy, BRS: Brugada syndrome, CPVT: catecholaminergic polymorphic ventricular tachycardia, D: definite, DCM: dilated cardiomyopathy, Dis: disputed, HCM: hypertrophic cardiomyopathy, L: limited, LQTS: long QT syndrome, M: moderate, NKDR: no known disease relationship, R: refuted, S: strong, SQTs: short QT syndrome.

### Variant Filtering

We retrieved the sequence for each gene in the candidate gene panel from gnomAD ^25^ v3.1 via the Google Cloud Platform BigQuery. We identified variants for each patient in the study cohort within the gene boundaries. Patient variants were merged to gnomAD. For variants present within gnomAD, we excluded those with minor allele frequency (MAF) > 0.5% (0.005). Patient variants were then subjected to analysis with the Ensembl Variant Effect Predictor (VEP) ^26^. For each variant, we utilized the Matched Annotation from the NCBI and EMBL-EBI (MANE) transcript to determine the predicted effect ^27^. We excluded variants with a predicted impact of “modifier” or “low” and included variants with a predicted impact of “moderate” or “high”.

### Variant Classification

Variants with impact of “moderate” or “high” in VEP (n=282) were further filtered by excluding missense variants with Combined Annotation-Dependent Depletion (CADD) score < 25 (n=222) ^28^ and those in TTN outside of the A-band (n=1), leaving 59 variants. The variant classification was performed with Sherloc and compared to variant classification within ClinVar ^29^. We modified Sherloc for use with gnomAD and ClinGen. For population data, we utilized a frequency of 0.01% rather than a count of 8 alleles^30^. For observations in affected individuals, we awarded 1 P point for variants in genes with a “definitive” or “strong” association with the phenotype in ClinGen. Criteria used for assigning points to each variant are detailed in Supplemental Table 1. Because variants 2-178650214-TTTTCCTCTTCAGGAGCAA-T and 19-49196501-TGCTGCGGGGGCC-T were each noted in 2 patients (patients 13/36 and patients 13/23, respectively) with differing phenotypes on autopsy (patient 13 SADS, patient 23 HCM, and patient 36 HCM), these 2 variants were included in both the SADS and HCM phenotype groups for variant-phenotype analysis (Figures 2 and 3). Because variants 2-178650214-TTTTCCTCTTCAGGAGCAA-T, 10-68199563-C-A, and 19-49196501-TGCTGCGGGGGCC-T were each noted in 2 patients, they were included separately for each ethnicity (Table 5).

**Figure 1.**
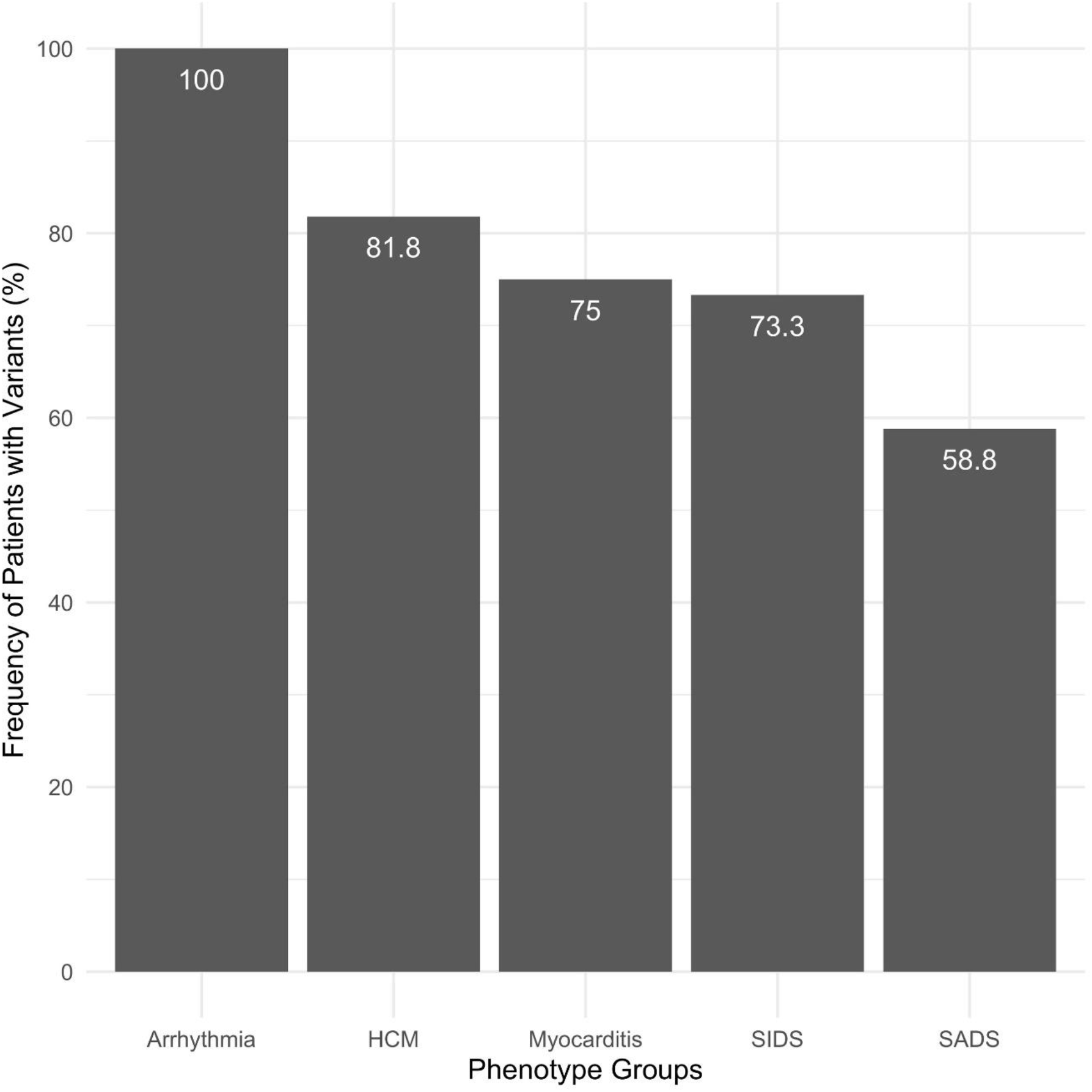
Comparison of the presence of gene variants (LB/VUS/LP/P) by phenotype group. We compared the presence of gene variants (LB/VUS/LP/P) to each patient’s phenotype group. In patients with a probable primary arrhythmia syndrome, all 5 patients (100%) possessed at least 1 variant. In patients with HCM, 9 of 11 patients (82%) possessed at least 1 variant. In patients with myocarditis, 3 of 4 patients (75%) possessed at least 1 variant. In 15 patients with SIDS, 11 patients (73%) possessed at least 1 variant. In 17 patients with SADS, 10 patients (59%) possessed at least 1 variant. HCM: hypertrophic cardiomyopathy, SADS: sudden arrhythmic death syndrome, SIDS: sudden infant death syndrome.

**Figure 2.**
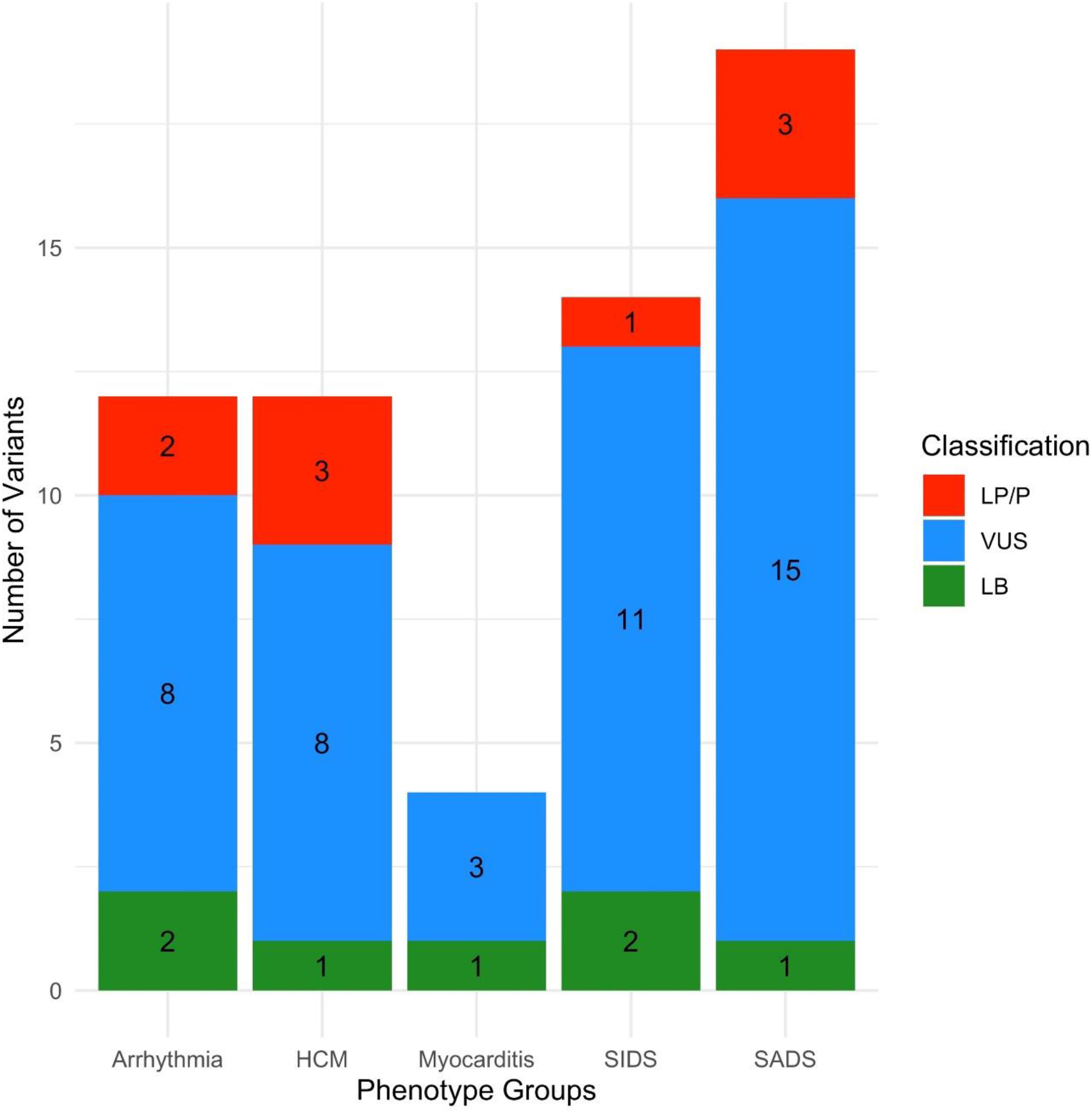
Comparison of gene variant classification (LB/VUS/LP/P) by phenotype group. We compared gene variant classification (LB/VUS/LP/P) to each patient’s phenotype group. In patients with a probable primary arrhythmia syndrome, 10 variants were classified as VUS/LP/P. In patients with HCM, 11 variants were classified as VUS/LP/P. In patients with myocarditis, 3 were classified as VUS. In patients with SIDS, 12 variants were classified as VUS/LP/P. In patients with SADS, 18 variants were classified as VUS/LP/P. HCM: hypertrophic cardiomyopathy, LP: likely pathogenic, P: pathogenic, SADS: sudden arrhythmic death syndrome, SIDS: sudden infant death syndrome, VUS=variant of uncertain significance.

**Figure 3.**
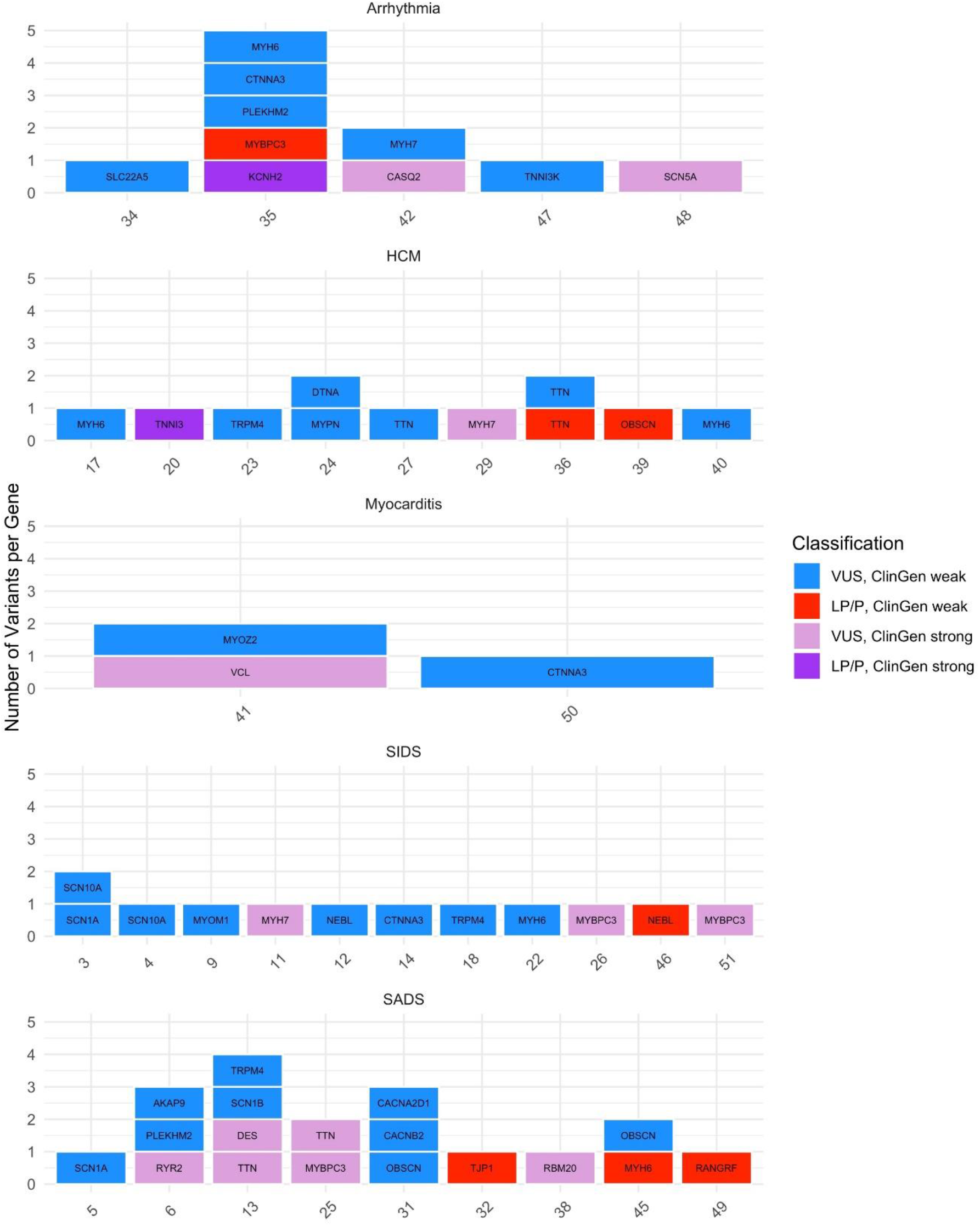
Comparison of genes with VUS/LP/P variants by phenotype group. We compared genes with VUS/LP/P variants by phenotype group. In this plot, each number on the x-axis represents a patient within that phenotype group, and each block represents that patient’s VUS/LP/P variants. Gene variants classified as VUS are colored blue if there was a weak association between that gene and the patient’s phenotype (according to the Clin-Gen gene-disease associations) and light purple if there was a strong association. Gene variants classified as LP/P are colored red if there was a weak association between that gene and the patient’s phenotype (according to the Clin-Gen gene-disease associations) and dark purple if there was a strong association.

The study protocol was approved by the institutional review boards of Ventura County Medical Center, Oregon Health and Science University, and the Cedars-Sinai Health System. This study includes both deceased and alive subjects. All analysis of deceased subjects is conducted in a de-identified manner. All subjects who are alive were approached and provided their consent to participate. There are no specific patient IDs in this study. Samples are numbered 1 through 52 and the IDs are not known to anyone outside the research group.

## Results

The study cohort included 52 SCA victims <21 years of age, of which 15 were <1 year of age. Overall, 45% of the cohort was female. The proportion of White victims was 67% (n=35), whereas 29% (n=15) were non-White, including 14% (n=7) Hispanic, 12% (n=6) Black, and 4% (n=2) Asian. We found that 40 (77%) patients in the study cohort died from their SCA, whereas 12 (23%) survived. Almost all deceased cases underwent autopsy (39/40). The most common phenotypes were SADS (32.7%, n=17), SIDS (29%, n=15), and HCM (21%, n=11), while 10% (n=5) had a probable primary arrhythmia syndrome and 8% (n=4) had myocarditis. We compared the demographics of the study cohort by survival and found that survivors were younger and more likely to have probable primary arrhythmia syndrome or SADS (Table 2).

**Table 2.**
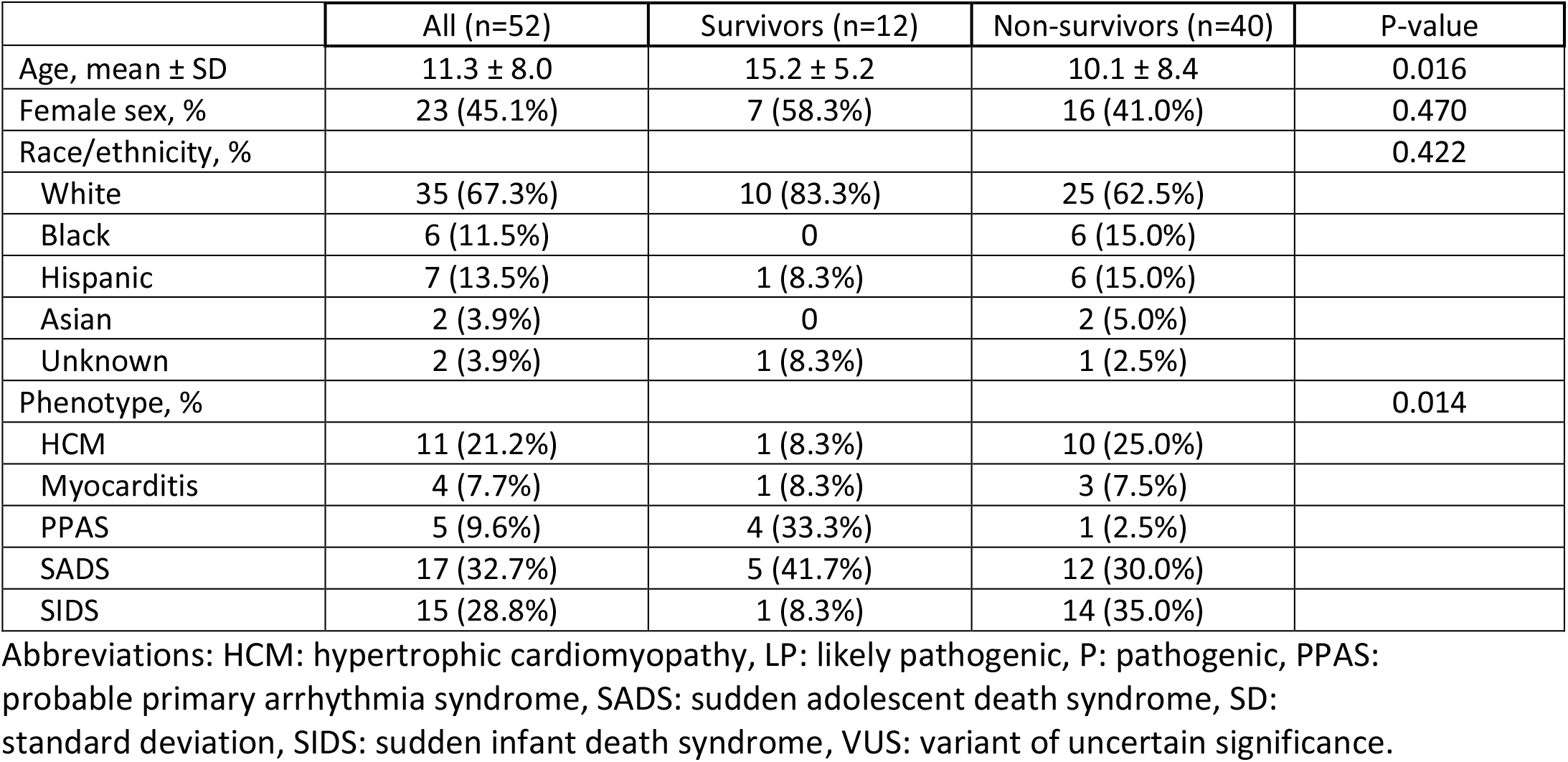
Demographic characteristics and phenotypes of the study subjects, in total and by survival after sudden cardiac arrest.

We identified 4,530,774 variants in 102 genes (genes KCNJ8 and TTR exhibited no variants) in 52 patients. This represented 266,145 unique variants, of which 96,665 (36%) were present in gnomAD and 169,480 (64%) were not present. We identified 282 variants with moderate/high effect which were then narrowed to 59 unique variants using stringent filtering criteria.

These 59 unique variants were in 32 genes (Table 3). The majority of variants, 40 of 59 (68%), were missense variants. In addition, there were 8 missense/splice region variants, 4 canonical splice site variants, 3 inframe deletions, 3 frameshifts and 1 stop gained. Of the variants, 39 had defined allelic frequencies in gnomAD and 20 did not. For those that had allelic frequencies in gnomAD, 31 (79%) were rare variants (frequency <0.1%). Variant effect predictions suggested loss of function for 8 variants: 4 frameshifts, 3 stop gained, and 4 canonical splice site variants. Classification via Sherloc indicated that 7 variants were likely benign (B), 43 of undetermined significance (VUS), 2 likely pathogenic (LP), and 7 pathogenic (P).

**Table 3.**
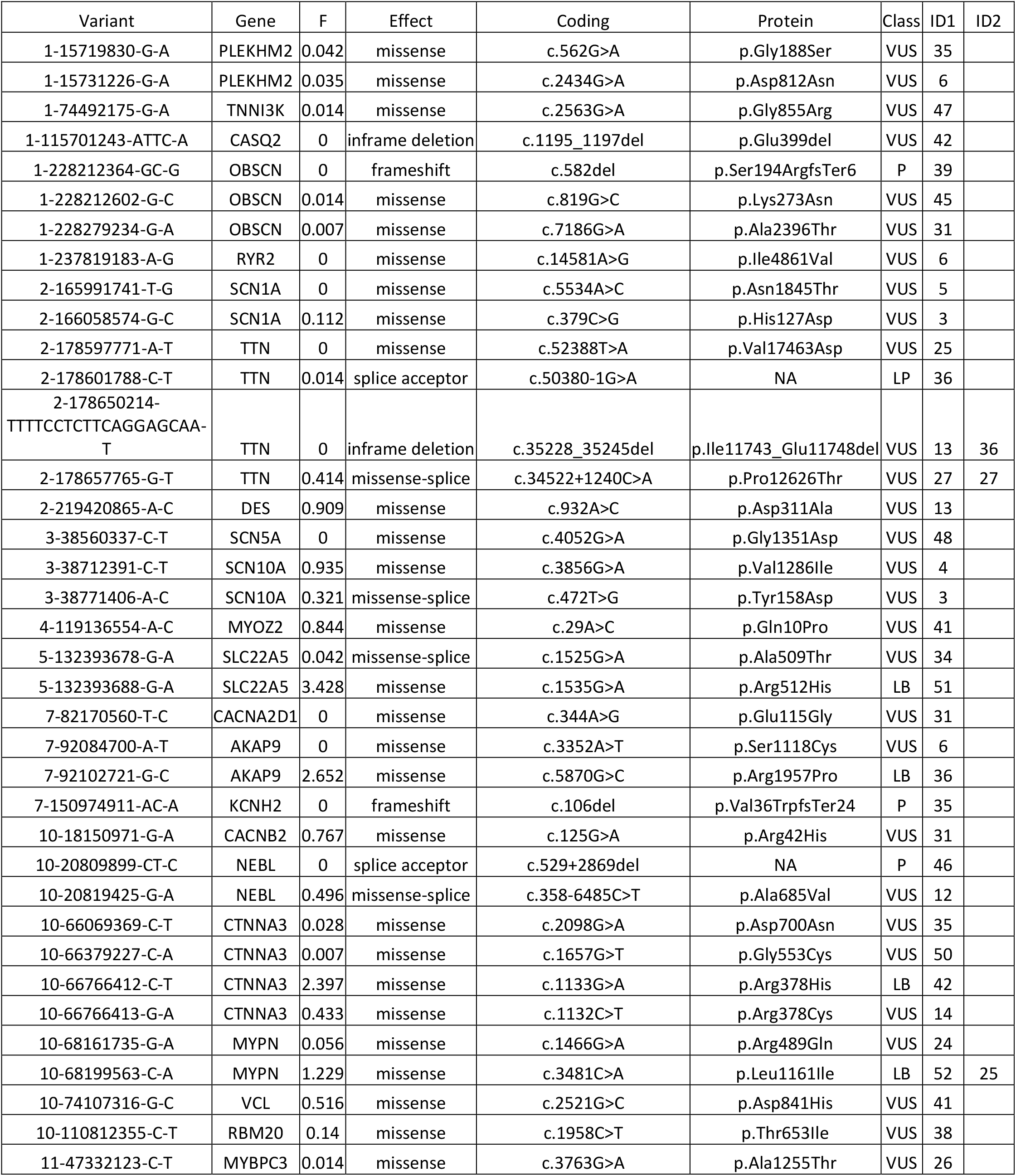
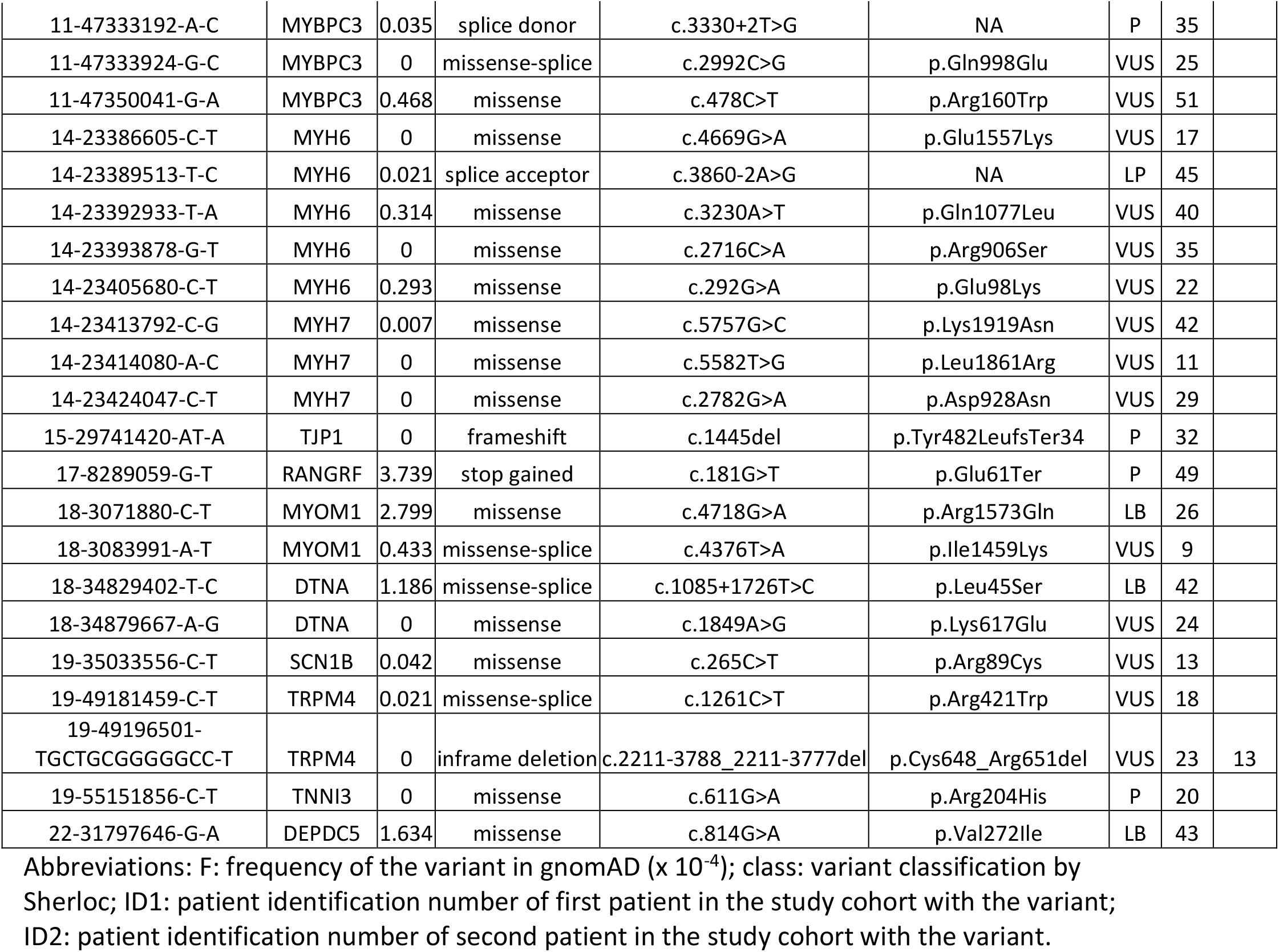
Variants detected in the present study.

Variant classification via Sherloc was compared to variant classifications within ClinVar (Table 4). For variants that were classified as B/LB by Sherloc, there was a strong correlation with ClinVar in that 5/7 (71%) were also B/LB in ClinVar. For variants that were classified as VUS by Sherloc, there was a fair correlation with ClinVar in that 13/43 (30%) were also VUS in ClinVar, but this was limited by 11/43 variants with conflicting evidence in ClinVar and another 11/43 variants missing from ClinVar. For variants that were classified as LP/P by Sherloc, there was a fair correlation with ClinVar in that 3/9 (30%) were also LP/P in ClinVar, again this was limited by 1/9 variants with conflicting evidence in ClinVar and another 3/9 variants missing from ClinVar. Two variants were classified as LP/P by Sherloc that were classified as B/LB in ClinVar.

**Table 4.** Variant classification via Sherloc compared to variant classifications within ClinVar

| Sherloc Class | Total Variants | ClinVar B/LB | ClinVar VUS | ClinVar LP/P | ClinVar Conflicting | ClinVar Missing |
| --- | --- | --- | --- | --- | --- | --- |
| B/LB | 7 | 5 | 0 | 0 | 2 | 0 |
| VUS | 43 | 7 | 13 | 1 | 11 | 11 |
| LP/P | 9 | 2 | 0 | 3 | 1 | 3 |

Abbreviations: B/LB: benign/likely benign, VUS: variant of undetermined significance, LP/P: likely pathogenic/pathogenic.

**Table 5.**
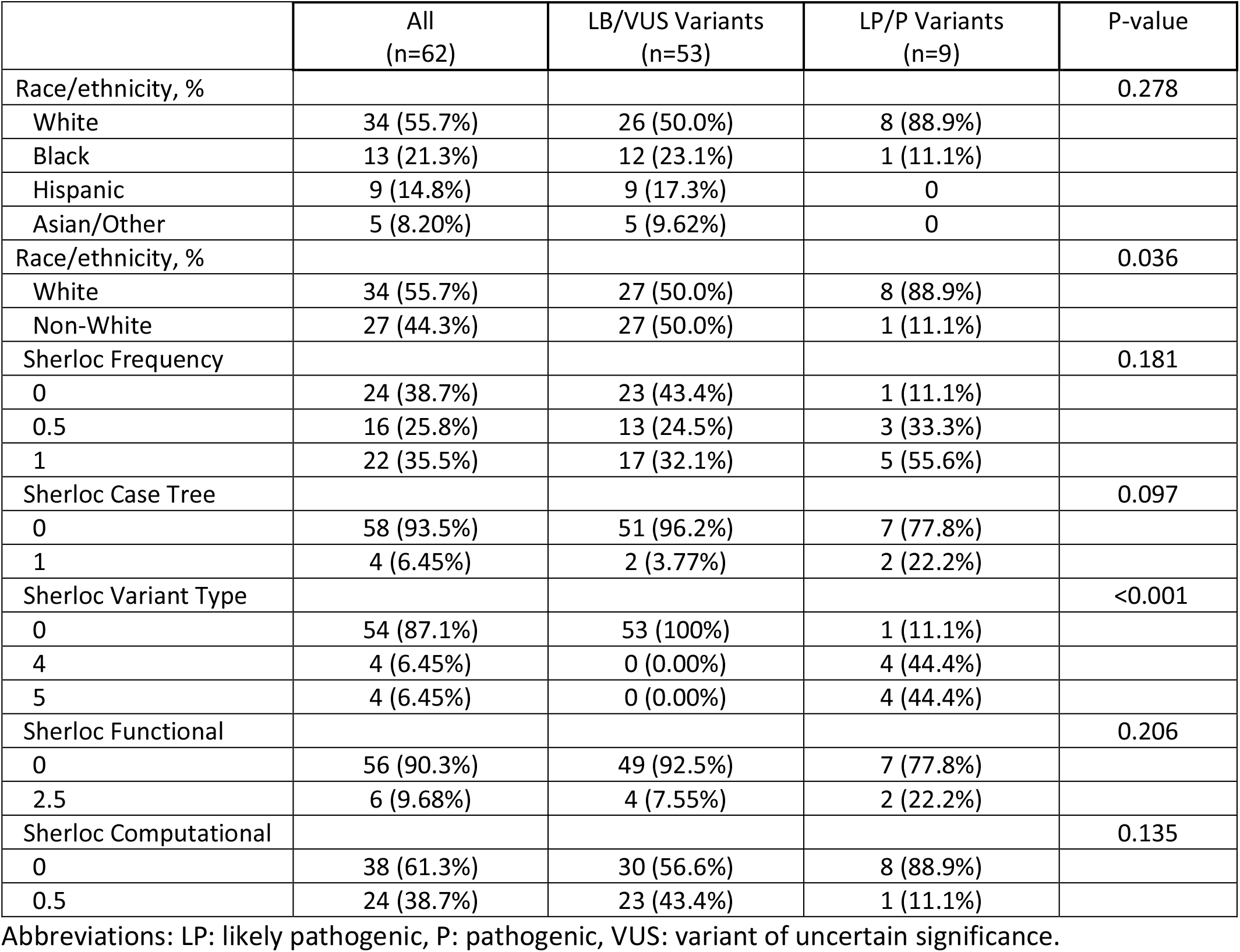
Comparison of frequency of LP/P variants by race/ethnicity of patient harboring the variant.

|  | All<br>(n=62) | LB/VUS Variants<br>(n=53) | LP/P Variants<br>(n=9) | P-value |
| --- | --- | --- | --- | --- |
| Race/ethnicity, % |  |  |  | 0.278 |
| White | 34 (55.7%) | 26 (50.0%) | 8 (88.9%) |  |
| Black | 13 (21.3%) | 12 (23.1%) | 1 (11.1%) |  |
| Hispanic | 9 (14.8%) | 9 (17.3%) | 0 |  |
| Asian/Other | 5 (8.20%) | 5 (9.62%) | 0 |  |
| Race/ethnicity, % |  |  |  | 0.036 |
| White | 34 (55.7%) | 27 (50.0%) | 8 (88.9%) |  |
| Non-White | 27 (44.3%) | 27 (50.0%) | 1 (11.1%) |  |
| Sherloc Frequency |  |  |  | 0.181 |
| 0 | 24 (38.7%) | 23 (43.4%) | 1 (11.1%) |  |
| 0.5 | 16 (25.8%) | 13 (24.5%) | 3 (33.3%) |  |
| 1 | 22 (35.5%) | 17 (32.1%) | 5 (55.6%) |  |
| Sherloc Case Tree |  |  |  | 0.097 |
| 0 | 58 (93.5%) | 51 (96.2%) | 7 (77.8%) |  |
| 1 | 4 (6.45%) | 2 (3.77%) | 2 (22.2%) |  |
| Sherloc Variant Type |  |  |  | <0.001 |
| 0 | 54 (87.1%) | 53 (100%) | 1 (11.1%) |  |
| 4 | 4 (6.45%) | 0 (0.00%) | 4 (44.4%) |  |
| 5 | 4 (6.45%) | 0 (0.00%) | 4 (44.4%) |  |
| Sherloc Functional |  |  |  | 0.206 |
| 0 | 56 (90.3%) | 49 (92.5%) | 7 (77.8%) |  |
| 2.5 | 6 (9.68%) | 4 (7.55%) | 2 (22.2%) |  |
| Sherloc Computational |  |  |  | 0.135 |
| 0 | 38 (61.3%) | 30 (56.6%) | 8 (88.9%) |  |
| 0.5 | 24 (38.7%) | 23 (43.4%) | 1 (11.1%) |  |

Abbreviations: LP: likely pathogenic, P: pathogenic, VUS: variant of uncertain significance.

Next, we assessed the pattern of the observed variants in the study cohort. We identified 38 patients with at least 1 variant. Of these, there were 24 patients with a single variant, 7 patients with 2 variants, 4 patients with 3 variants, 2 patients with 4 variants, and 1 patient with 5 variants. All variants were heterozygous, except for TTN variant 2-178657765-G-T observed in patient 27, which was homozygous. Three variants were present in more than 1 patient in the cohort: (1) TTN variant 2-178650214-TTTTCCTCTTCAGGAGCAA-T was present in patients 13 and 26 and was classified as VUS, (2) TRPM4 variant 19-49196501-TGCTGCGGGGGCC-T was present in patients 13 and 23 and was classified as VUS, and (3) MYPN variant 10-68199563-C-A was present in patients 25 and 52 and was classified as LB.

We then compared the presence of gene variants (LB/VUS/LP/P) to each patient’s phenotype group (Figure 1). Overall, 38 of 52 patients (73%) possessed at least 1 variant. In patients with a probable primary arrhythmia syndrome, all 5 patients possessed at least 1 variant, with all 5 patients (100% of the group) possessing at least 1 VUS/LP/P variant. In patients with HCM, 9 of 11 patients possessed at least 1 variant, with these 9 patients (82% of the group) possessing at least 1 VUS/LP/P variant. In patients with myocarditis, 3 of 4 patients possessed at least 1 variant, with 2 patients (50% of the group) possessing at least 1 VUS/LP/P. Of 15 patients with SIDS, 11 patients possessed at least 1 variant, with these 11 patients (73% of the group) possessing at least 1 VUS/LP/P variant. Of 17 patients with SADS, 10 patients possessed at least 1 variant, with 9 patients (53% of the group) possessing at least 1 VUS/LP/P variant.

We then compared the gene variant classification (LB/VUS/LP/P) to each patient’s phenotype group (Figure 2). In patients with probable primary arrhythmia syndrome, there were a total of 12 variants, with 2 classified as LB, 8 classified as VUS, and 2 classified as LP/P. In patients with HCM, there were a total of 12 variants, with 1 classified as LB, 8 classified as VUS, and 3 classified as LP/P. In patients with myocarditis, there were a total of 4 variants, with 1 classified as LB, 3 classified as VUS, and none classified as LP/P. In patients with SIDS, there were a total of 14 variants, with 2 classified as LB, 11 classified as VUS, and 1 classified as LP/P. In patients with SADS, there were a total of 19 variants, with 1 classified as LB, 15 classified as VUS, and 3 classified as LP/P.

In total we identified 38 VUS/LP/P variants in genes associated with cardiomyopathic conditions (ARVC, DCM, or HCM) at any strength of gene-disease association in the ClinGen panel (Table 1). Of these 38 variants, 2 variants (variant 3-38560337-C-T in SCN5A and 1-237819183-A-G in RYR2) were also associated with arrhythmic conditions. We identified 12 VUS/LP/P variants in genes associated only with arrhythmic conditions (BRS, CPVT, LQTS, or SQTS) at any strength of gene-disease association in the ClinGen panel. Finally, we identified 2 VUS/LP/P variants in genes associated with sudden death in epilepsy.

We then compared the genes harboring VUS/LP/P variants by phenotype group (Figure 3). In patients with a probable primary arrhythmia syndrome with VUS/LP/P variants, 3 were in genes definitively/strongly/moderately associated with primary arrhythmia syndromes in ClinGen, specifically CASQ2 (associated with CPVT), KCNH2 (associated with LQTS), and SCN5A (associated with BrS). In patients with HCM with VUS/LP/P variants, 2 were in genes definitively/strongly/moderately associated with HCM, specifically MYH7 and TNNI3. In patients with myocarditis with VUS/LP/P variants, 1 was in a gene definitively/strongly/moderately associated with DCM, specifically VCL. Although there is no ClinGen panel for SIDS, 3 variants were in genes definitively/strongly/moderately associated with phenotypes in the panel, 2 variants in MYBPC3 (associated with HCM), and 1 variant in MYH7 (associated with DCM). Finally, while there is no ClinGen panel for SADS, 6 variants were in genes definitively/strongly/moderately associated with phenotypes in the panel, specifically DES (associated with DCM), MYBPC3 (associated with HCM), RBM20 (associated with DCM), RYR2 (associated with CPVT), and 2 TTN variants (associated with DCM).

Lastly, we compared characteristics of variants by their classification as LB/VUS versus LP/P (Table 5). We found no significant difference between the frequency of LB/VUS versus LP/P variants by ethnic group (White, Black, Hispanic, Asian/Other/Unknown), however when variants were stratified as occurring in White or non-White patients, we found significantly more LP/P variants in White (8/9 88.9%) as compared to non-White patients (1/9 11.1%, p = 0.036). We then compared the Sherloc pathogenicity criteria of variants between these groups. We found no significant differences for the frequency, case tree, functional or computational criteria, but did find a significantly higher frequency of loss of function and canonical splice site variants in the LP/P variants.

## Discussion

Here we have studied the prevalence of rare genetic variants in young victims of SCA residing in 2 US communities. We found that the prevalence of potentially disease-associated rare variants was 69% as 36 of 52 patients possessed at least 1 potentially disease-related variant. Patients with clinical histories suggesting a primary arrhythmia syndrome or HCM were most likely to have a clinically significant variant, with 100% and 82% of these groups, respectively, possessing at least 1 P/LP/VUS variant. Patients with clinical histories suggesting myocarditis, SIDS, or SADS were less likely to harbor a genetic variant within the 104 gene panel that was used. The ability to analyze SCA-related variation at the population level (as opposed to referral samples) is a strength of this study. In addition, unlike most published studies that have focused on SCD, were also able to study survivors of SCA (23% in our cohort). Since survival from SCA is largely a function of time to resuscitation, inclusion of this subset provides a more comprehensive analysis.

Although we identified a high frequency of potentially disease-related rare variants (i.e. VUS, LP, or P variants), we identified 8 of 52 patients (15%) with an actionable LP/P variant. The prevalence of actionable variants was somewhat lower in our study than in the study by Bagnall et. al. where they identified 27% of unexplained young SCD cases with an LP/P variant ^6^. In addition, studies from Canadian pediatric centers, Catalonia, and Denmark have also reported higher (30-40%) proportions of positive findings^31-33^. In contrast, several other studies have reported clinically actionable variants in 10-18% of young SCD cases^3, 8, 24, 34-36^. Regarding SIDS cases, our results are consistent with the study by Tester et al^37^ which reported that 4.3% of 419 SIDS cases harbored an LP/P variant (1/15=6.7% in our study). Direct comparison to previous studies is complicated due to differences in e.g., SCD/SCA phenotypes, gene panels, and variant interpretation strategies, but given that previous studies report a positive result in 10 to 30% of young SCD cases, our results are within the lower range of this estimate. While the exact reason is not fully clear, we hypothesize that previously reported series, that have mainly utilized data obtained from special referral clinics or registries, may be prone to a selection bias that leads to a higher observed prevalence of gene variants.

Importantly, we found that clinically actionable LP/P variants were less likely to be identified in non-White patients within the study cohort. Furthermore, this difference was largely due to a significantly lower frequency of loss of function and canonical splice site variants. Black patients with cardiomyopathy and SCA have previously been found to harbor a lower prevalence of pathogenic variants^3, 38-40^, but reasons for this lower prevalence were not rigorously investigated as we have done in this study. Previously Hagerty et al. found that titin truncating variants were associated with DCM in White but not in Black patients^41^. On the other hand, the incidence of SCA is similar in White and Black patients in the community^11^. Taken together, these results suggest that rare genetic variants associated with SCA may differ by race/ethnicity and that additional methods such as transcriptome sequencing^42^ and molecular assays of variant effect^43^ may be important to help classify the pathogenicity of variants identified in non-White victims of SCA.

Our results describe a higher proportion of cardiomyopathy-related variants than ion channel gene variants. Although variants in cardiomyopathy-related genes are usually present in SCD cases due to inherited cardiomyopathies, such variants have also been associated with nonspecific subtle structural alterations (*e.g*. myocardial fibrosis)^36, 44, 45^, and structurally normal hearts^46^. It has been hypothesized that variants in cardiomyopathy-related genes may contribute to SCA even without overt clinical signs of inherited cardiomyopathy, thus representing “concealed cardiomyopathy”. However, the clinical significance of subtle non-specific myocardial alterations is not fully clear, and more work is needed to study the wide clinical spectrum of inherited cardiomyopathies to recognize patients that are at a high risk of future arrhythmias.

Despite the relatively small number of clinically actionable gene variants, our findings are consistent with current guidelines ^47^ which support the use of genetic testing in young SCA victims. Cascade genetic testing facilitates recognition of family members that are at an increased risk of SCD and may therefore benefit from preventive interventions. Family history of SCD is also a risk factor for SCD ^48^, and hence family members with positive genetic testing may be at a significantly increased risk of SCD and require regular follow-up. Sudden and unexpected death in a young person may cause a significant amount of uncertainty and anxiety in family members, and negative test results also have important clinical implications as those family members can be discharged from follow-up and unnecessary preventive interventions.

The possibility of genetic screening to prevent SCD in targeted young populations (*e.g*. athletes) has also been discussed, but the effectiveness of such an approach is yet to be investigated^49^. Although our study demonstrated that a notable proportion of young SCA cases may harbor potentially disease-associated rare gene variants (VUS/LP/P) since only 15% of variants were LP/P it remains unclear whether primary screening would be effective due to the high frequency of non-LP/P variants. SCD risk is highly variable in patients with inherited cardiac disease and usually cannot be determined based on single gene variants. Incomplete penetrance and variable phenotypic expression suggest that the inheritance pattern is not fully monogenic and is likely affected by modifying gene variants and environmental factors as well. Hence, the penetrance of inherited cardiac diseases is likely to be dependent on the genomic context: gene-positive patients with a family history of inherited cardiac disease may express higher penetrance, while the penetrance of inherited cardiac diseases is possibly lower in the general screening population^50, 51^.

Our study has several novel aspects. First, the study cohort has been drawn from 2 community-based SCA cohorts. Given that SCA in the young is a rare event, collecting such cohorts has required a long effort in identifying and adjudicating possible SCA cases. Another strength of our study is well-adjudicated phenotypes, as almost all the non-survivors underwent autopsy (39/40, 97.5%). Next, we utilized an evidence-based approach using ClinGen panels to select genes for study. Lastly, our study cohort was racially/ethnically diverse in that 15 of 50 (30%) patients in the study cohort and 27 of 62 (44%) variants were in non-White individuals. This makes our study cohort one of the most diverse cohorts of young patients with SCA.

Our study also has some limitations that should be considered. At first, our variant interpretation strategy favors specificity over sensitivity, and therefore, we may have missed some disease-associated variants in the process. Secondly, we did not have a control group and our study also lacks segregation data, which limits the interpretation of variant pathogenicity, especially for VUS. Thirdly, given that our gene panel included only 104 genes related to inherited cardiac disorders, we cannot exclude the possibility that some of our patients may have also harbored disease-causing variants beyond our gene panel.

## Conclusions

In this community-based study of young SCA victims, we found that the prevalence of potentially disease-related variants was 69% and the prevalence of pathogenic/likely pathogenic variants was 15%. As expected, SCAs that were likely due to primary arrhythmic disorders harbored variants in genes related to CPVT, LQTS, and BrS, while a wide spectrum of SCAs due to HCM, SADS, or myocarditis harbored potentially disease-associated variants in cardiomyopathy related genes. These findings suggest that the proportion of clinically actionable variants at the community level may be somewhat lower than previously anticipated. However, the overall prevalence of likely pathogenic or uncertain variants highlights the need for further efforts to improve the diagnostic yield of young SCA cases with a strong suspicion of genetic cardiac disease.

## Data Availability

The data that support the findings of this study are available from the corresponding author upon reasonable request.

## Acknowledgments

The authors gratefully acknowledge the contributions of the Portland OR and Ventura CA communities as well as their EMS, health and medical examiner systems, without whom this work could not be accomplished.

