## Supplemental Table 1 for "Rare Genetic Variants Associated with Sudden Cardiac Arrest in the Young: A Prospective, Population-Based Study"

**Supplemental Table.** Points awarded to each variant by application of the Sherloc algorithm.

| Variant | Frequency Pathogenic | Frequency Benign | Case Tree | Variant Type | Functional | Computational | Sum Pathogenic | Sum Benign | Assignment | ID1 | ID2 |
| --- | --- | --- | --- | --- | --- | --- | --- | --- | --- | --- | --- |
| 2-166058574-G-C | 0 | 1 | 0 | 0 | 0 | 0.5 | 0.5 | 1 | VUS | 3 |  |
| 3-38771406-A-C | 0 | 1 | 0 | 0 | 2.5 | 0.5 | 3 | 1 | VUS | 3 |  |
| 3-38712391-C-T | 0 | 1 | 0 | 0 | 0 | 0.5 | 0.5 | 1 | VUS | 4 |  |
| 2-165991741-T-G | 1 | 0 | 0 | 0 | 0 | 0.5 | 1.5 | 0 | VUS | 5 |  |
| 1-15731226-G-A | 0.5 | 0 | 0 | 0 | 0 | 0.5 | 1 | 0 | VUS | 6 |  |
| 1-237819183-A-G | 1 | 0 | 0 | 0 | 0 | 0.5 | 1.5 | 0 | VUS | 6 |  |
| 7-92084700-A-T | 1 | 0 | 0 | 0 | 0 | 0 | 1 | 0 | VUS | 6 |  |
| 18-3083991-A-T | 0 | 1 | 0 | 0 | 0 | 0 | 0 | 1 | VUS | 9 |  |
| 14-23414080-A-C | 1 | 0 | 0 | 0 | 0 | 0 | 1 | 0 | VUS | 11 |  |
| 10-20819425-G-A | 0 | 1 | 0 | 0 | 0 | 0 | 0 | 1 | VUS | 12 |  |
| 19-35033556-C-T | 0.5 | 0 | 0 | 0 | 0 | 0.5 | 1 | 0 | VUS | 13 |  |
| 19-49196501-TGCTGCGGGGGCC-T | 1 | 0 | 0 | 0 | 0 | 0 | 1 | 0 | VUS | 13 | 23 |
| 2-178650214-TTTTCCTCTTCAGGAGCAA-T | 1 | 0 | 0 | 0 | 0 | 0 | 1 | 0 | VUS | 13 | 36 |
| 2-219420865-A-C | 0 | 1 | 0 | 0 | 2.5 | 0.5 | 3 | 1 | VUS | 13 |  |
| 10-66766413-G-A | 0 | 1 | 0 | 0 | 0 | 0 | 0 | 1 | VUS | 14 |  |
| 14-23386605-C-T | 1 | 0 | 0 | 0 | 0 | 0 | 1 | 0 | VUS | 17 |  |
| 19-49181459-C-T | 0.5 | 0 | 0 | 0 | 0 | 0.5 | 1 | 0 | VUS | 18 |  |
| 19-55151856-C-T | 1 | 0 | 1 | 0 | 2.5 | 0.5 | 5 | 0 | P | 20 |  |
| 14-23405680-C-T | 0 | 1 | 0 | 0 | 0 | 0 | 0 | 1 | VUS | 22 |  |
| 10-68161735-G-A | 0.5 | 0 | 0 | 0 | 0 | 0 | 0.5 | 0 | VUS | 24 |  |
| 18-34879667-A-G | 1 | 0 | 0 | 0 | 0 | 0 | 1 | 0 | VUS | 24 |  |
| 10-68199563-C-A | 0 | 3 | 0 | 0 | 0 | 0.5 | 0.5 | 3 | LB | 25 | 52 |
| 11-47333924-G-C | 1 | 0 | 0 | 0 | 0 | 0 | 1 | 0 | VUS | 25 |  |
| 2-178597771-A-T | 1 | 0 | 0 | 0 | 0 | 0 | 1 | 0 | VUS | 25 |  |
| 11-47332123-C-T | 0.5 | 0 | 0 | 0 | 0 | 0.5 | 1 | 0 | VUS | 26 |  |
| 18-3071880-C-T | 0 | 3 | 0 | 0 | 0 | 0.5 | 0.5 | 3 | LB | 26 |  |
| 2-178657765-G-T | 0 | 1 | 0 | 0 | 0 | 0 | 0 | 1 | VUS | 27 | 27 |
| 14-23424047-C-T | 1 | 0 | 1 | 0 | 0 | 0 | 2 | 0 | VUS | 29 |  |
| 1-228279234-G-A | 0.5 | 0 | 0 | 0 | 0 | 0 | 0.5 | 0 | VUS | 31 |  |
| 10-18150971-G-A | 0 | 1 | 0 | 0 | 0 | 0 | 0 | 1 | VUS | 31 |  |
| 7-82170560-T-C | 1 | 0 | 0 | 0 | 0 | 0 | 1 | 0 | VUS | 31 |  |
| 15-29741420-AT-A | 1 | 0 | 0 | 5 | 0 | 0 | 6 | 0 | P | 32 |  |
| 5-132393678-G-A | 0.5 | 0 | 0 | 0 | 0 | 0 | 0.5 | 0 | VUS | 34 |  |
| 1-15719830-G-A | 0.5 | 0 | 0 | 0 | 0 | 0 | 0.5 | 0 | VUS | 35 |  |
| 10-66069369-C-T | 0.5 | 0 | 0 | 0 | 0 | 0.5 | 1 | 0 | VUS | 35 |  |
| 11-47333192-A-C | 0.5 | 0 | 0 | 4 | 2.5 | 0 | 7 | 0 | P | 35 |  |
| 14-23393878-G-T | 1 | 0 | 0 | 0 | 0 | 0 | 1 | 0 | VUS | 35 |  |
| 7-150974911-AC-A | 1 | 0 | 1 | 5 | 0 | 0 | 7 | 0 | P | 35 |  |
| 2-178601788-C-T | 0.5 | 0 | 0 | 4 | 0 | 0 | 4.5 | 0 | LP | 36 |  |
| 7-92102721-G-C | 0 | 3 | 0 | 0 | 0 | 0.5 | 0.5 | 3 | LB | 36 |  |
| 10-110812355-C-T | 0 | 1 | 0 | 0 | 0 | 0.5 | 0.5 | 1 | VUS | 38 |  |
| 1-228212364-GC-G | 1 | 0 | 0 | 5 | 0 | 0 | 6 | 0 | P | 39 |  |
| 14-23392933-T-A | 0 | 1 | 0 | 0 | 0 | 0.5 | 0.5 | 1 | VUS | 40 |  |
| 10-74107316-G-C | 0 | 1 | 0 | 0 | 2.5 | 0 | 2.5 | 1 | VUS | 41 |  |
| 4-119136554-A-C | 0 | 1 | 0 | 0 | 0 | 0 | 0 | 1 | VUS | 41 |  |
| 1-115701243-ATTC-A | 1 | 0 | 0 | 0 | 0 | 0 | 1 | 0 | VUS | 42 |  |
| 10-66766412-C-T | 0 | 3 | 0 | 0 | 0 | 0.5 | 0.5 | 3 | LB | 42 |  |
| 14-23413792-C-G | 0.5 | 0 | 0 | 0 | 0 | 0.5 | 1 | 0 | VUS | 42 |  |
| 18-34829402-T-C | 0 | 3 | 0 | 0 | 0 | 0 | 0 | 3 | LB | 42 |  |
| 22-31797646-G-A | 0 | 3 | 0 | 0 | 0 | 0.5 | 0.5 | 3 | LB | 43 |  |
| 1-228212602-G-C | 0.5 | 0 | 0 | 0 | 0 | 0 | 0.5 | 0 | VUS | 45 |  |
| 14-23389513-T-C | 0.5 | 0 | 0 | 4 | 0 | 0 | 4.5 | 0 | LP | 45 |  |
| 10-20809899-CT-C | 1 | 0 | 0 | 4 | 0 | 0 | 5 | 0 | P | 46 |  |
| 1-74492175-G-A | 0.5 | 0 | 0 | 0 | 0 | 0 | 0.5 | 0 | VUS | 47 |  |
| 3-38560337-C-T | 1 | 0 | 1 | 0 | 0 | 0.5 | 2.5 | 0 | VUS | 48 |  |
| 17-8289059-G-T | 0 | 3 | 0 | 5 | 0 | 0 | 5 | 3 | P | 49 |  |
| 10-66379227-C-A | 0.5 | 0 | 0 | 0 | 0 | 0.5 | 1 | 0 | VUS | 50 |  |
| 11-47350041-G-A | 0 | 1 | 0 | 0 | 0 | 0.5 | 0.5 | 1 | VUS | 51 |  |
| 5-132393688-G-A | 0 | 3 | 0 | 0 | 2.5 | 0 | 2.5 | 3 | LB | 51 |  |
